# Service disruptions and stockouts: Ongoing impacts of U.S. funding freezes on HIV care across clinics in 38 low- and middle-income countries in late 2025

**DOI:** 10.64898/2026.08.04.26359701

**Authors:** Ellen Brazier, Maabo Kludze, Fernanda Maruri, Annabelle Niyongabo, Philip Kreniske, Stephany N. Duda, Denis Nash

**Affiliations:** Institute for Implementation Science in Population Health, City University of New York, Graduate School of Public Health and Health Policy, New York, NY, United States; City University of New York, Graduate School of Public Health and Health Policy, New York, NY, United States; Division of Infectious Diseases, Department of Medicine, Vanderbilt University Medical Center, Nashville, TN, United States; Association Nationale de Soutien aux Séropositifs et malades du sida-Santé PLUS (ANSS-Santé PLUS), Bujumbura, Burundi; Department of Biomedical Informatics, Vanderbilt University Medical Center, Nashville, TN, United States

## Abstract

**Introduction:** After U.S. foreign assistance was frozen in January 2025, empirical data on the status of the HIV response has been limited. To better understand the ongoing impacts of changes in U.S. foreign assistance, we launched an open survey to assess disruptions in HIV-related care among clinics and programs in low- and middle-income countries (LMICs).

**Methods:** Conducted from August to December 2025, the survey explored U.S. foreign assistance-related disruptions in HIV service delivery, medication availability, laboratory services and clinic operations; whether disruptions were fully resolved at the survey timepoint; and the introduction of clinic mitigation strategies. Data on other impacts of U.S. funding changes were explored through an open-ended question. A convergent mixed-methods design, involving parallel quantitative and qualitative analyses and merging of findings from each, was used to examine the impacts of U.S. funding freezes on HIV-related care.

**Results:** We received 158 responses from 38 LMICs, including 30 countries supported by the U.S. President’s Emergency Fund for AIDS Relief (PEPFAR) at the beginning of 2025 (n=123 responses) and eight non-PEPFAR countries (n=35 responses). Respondents represented health centers (25%), hospitals (31%), dedicated HIV clinics and drop-in centers (39%), and multi-site programs (4%), with a majority (59%) in the non-governmental/private sector. Overall, 81% reported disruptions in at least one HIV-related service since January 2025 because of changes in U.S. foreign assistance, with most also reporting disruptions in medication availability, laboratory services, and clinic operations. The largest reported disruptions were in the areas of pre-exposure prophylaxis (67%) and HIV testing (63%), along with patient tracing (67%), adherence support (63%) and services to key populations (64%). Disruptions were more prevalent in PEPFAR-supported countries and were more likely to be “not fully resolved” by time of survey completion. Qualitative data highlighted the impact of U.S. foreign assistance disruptions on the erosion of client trust in the health system and strains on staff morale.

**Conclusions:** Substantial and sustained disruptions in HIV prevention and care reported by diverse clinics in LMICs reinforce concerns that recent funding shifts could reverse progress in ending the HIV epidemic, particularly for vulnerable and key populations.

## I. Introduction

In January 2025, the U.S. government issued a series of executive orders, including an immediate, 90-day pause of all U.S. foreign assistance for a review and affirmative determination that each program and obligation was aligned with the administration’s policies and priorities [1, 2]. In the following days, the U.S. Agency for International Development (USAID), an agency responsible for obligating and implementing 60% of U.S. funding for the U.S. President’s Emergency Fund for AIDS Relief (PEPFAR) [3], issued stop work orders for existing awards, suspending activities related to all U.S.-funded global health programs. While limited waivers were issued in early February 2025 for PEPFAR and other global health programs to resume “urgent life-saving HIV treatment services” (e.g., HIV treatment, prevention of mother-to child or vertical transmission (PVT)) and other life-saving services (e.g., tuberculosis, malaria, treatment of severe acute malnutrition and other life-threatening conditions), there were delays in authorizing the reinstatement of services covered by these waivers, and by early March 2025, 83% of USAID programs were cancelled [1].

Initial efforts to document the impact of U.S. foreign assistance freezes indicated that uncertainty about funding, suspended payments and the cancelation of awards to PEPFAR and USAID implementing partners resulted in clinic closures, staff layoffs, interruptions in HIV-related service delivery and shortages of antiretroviral medications (ARVs) in the first quarter of 2025 [4–9]. Subsequent empirical stock-taking efforts confirmed the substantial scale of disruptions in HIV-related services; a study in KwaZulu-Natal, South Africa found that 42% of clinics surveyed had experienced disruptions in services, operations or staffing interruptions because of U.S. assistance cuts [10]. Similarly, a multiregional survey of clinics and programs across 32 countries participating in the International epidemiology Databases to Evaluate AIDS (IeDEA) found that almost half of responding sites had experienced disruptions in HIV-related services because of U.S. assistance freezes, with most reporting that all disruptions were not fully resolved by June-July 2025 [11].

To provide a broader understanding of the ongoing impacts of changes in U.S. foreign assistance and contextualize findings from our earlier survey of clinics and programs participating in IeDEA collaboration, we launched an open survey among HIV clinics and programs in low- and middle-income countries, including countries supported by PEPFAR at the start of 2025, as well as countries without PEPFAR support.

## II. Methods

### Study design and data

This study employed a convergent mixed-methods design[12], involving parallel analysis of qualitative and quantitative data and merging of findings from each, to assess the impacts of U.S. foreign assistance freezes on HIV-related care at participating clinics.

Data were obtained through an open cross-sectional survey aimed at HIV clinics and programs in low- and middle-income countries. The questionnaire was based on an earlier survey conducted among clinics participating in the IeDEA research collaboration [11]. Questions explored whether selected HIV-related services, medication supplies, laboratory services and clinic operations (Table 1) had been disrupted or suspended since January 2025 because of changes in U.S. government policy or funding, as well as whether certain mitigation strategies had been introduced in response to funding disruptions. For each service-related disruption, response options included “Yes”, “No”, “Do not know” and “Not applicable” for services not provided at the clinic. For any disruptions attributed to U.S. assistance freezes, respondents were further asked whether the disruption was partially or fully resolved at the time of survey completion. For each mitigation measure introduced in response to funding disruptions, respondents were asked to indicate whether or not the measure was still in effect.

**Table 1.** Survey domains.

|  |  |
| --- | --- |
| **HIV-related services** | HIV counseling and testing<br>HIV treatment<br>Pediatric HIV services<br>PVT services<br>TB/MDR-TB treatment<br>PrEP consultations<br>PEP consultations<br>Sexual/gender-based violence services<br>Condoms<br>Contraceptives/family planning services |
| **Medication availability** | Adult antiretrovirals (ARVs)<br>Pediatric ARVs<br>Cotrimoxazole (Bactrim, Septra, Trimethoprim Sulfamethoxazole)<br>Anti-TB medications<br>Contraceptives<br>Anti-malarials |
| **Laboratory services** | HIV diagnosis/confirmatory testing<br>Viral load testing<br>Early infant diagnosis<br>HIV drug resistance testing<br>CD4 cell count testing<br>TB diagnosis |
| **Clinic operations** | Clinic closure(s) or reduced hours/days of operation<br>Staffing shortages or layoffs<br>Salary reductions for HIV care providers<br>Multi-month dispensing of medication<br>Processing of lab specimens<br>Adherence support programs<br>Community-based activities<br>Patient tracing activities (i.e., patients lost to follow-up)<br>TB contact tracing<br>Services for key populations<br>Services for orphans/vulnerable children<br>Patient record management |
| **Mitigation strategies** | Introduction/expansion of MMD for eligible patients<br>Introduction/expansion of cost-sharing measures<br>Advocacy, fundraising or new partnership(s) to support service provision<br>Introduction/expansion of telemedicine<br>Patient referral/transfer to other clinics |

**Abbreviations:** ARVs: antiretrovirals; MMD: Multi-month dispensing; PEP: Post-exposure prophylaxis; PrEP: Pre-exposure prophylaxis; PVT: Prevention of vertical transmission; TB/MDR-TB: Tuberculosis or multi-drug-resistant tuberculosis.

The questionnaire also included questions about the respondent’s clinic or program, including the country and city/municipality where the facility was located; facility type (e.g., health center, district hospital, referral or teaching hospital, or other); facility administration (e.g., public/government sector vs. private/non-governmental); population served (e.g., predominantly urban, predominantly rural or mixed), number of people living with HIV (PLHIV) served (e.g., <50, 51-100, 101-200, 201-500, 501-1000, 1000+); and age-group(s) served (e.g., children 0-9 years, adolescents 10-19 years, young adults 20-24 years and adults 25+). The survey also included a question on respondents’ role (e.g., clinical director/staff vs. other HIV program staff) and an optional open-ended question on other impacts of U.S. policy and funding changes at their clinic. While the survey was answered anonymously, respondents were asked to provide the name of their facility solely for the purpose of identifying duplicate responses from the same clinic.

The survey was programmed in REDCap [13, 14], and REDCap’s multi-language module was used to make the questionnaire available in English, French, Spanish and Swahili. Designated as non-human subjects research per 45 CFR §46.102(l) by Vanderbilt University Medical Center’s IRB (#250988) not requiring informed consent, the survey was launched on August 4, 2025 and closed on December 31, 2025. Information about the survey and an open survey link were shared via personal outreach to individuals affiliated with various multilateral, international and national organizations involved in the HIV programs, including UN agencies, PEPFAR implementing partners and local and international non-governmental organizations operating or managing multiple clinics within their respective countries. Additionally, a description of the survey and survey link were shared with over 10,000 active members of IAS – the International AIDS Society – in over 170 countries via newsletter updates in September and November 2025.

Survey data were linked to publicly available data related to U.S. foreign assistance for each country [15], along with each country’s annual PEPFAR budget allocation [16] and disbursements from the Global Fund to Fight AIDS, Tuberculosis and Malaria (hereafter Global Fund) [17], Based on PEPFAR budget allocation data for 2025, countries were categorized as being PEPFAR-supported or non-PEPFAR-supported, and non-PEPFAR-supported countries were further categorized as Global Fund-supported if they had a Global Fund disbursement in 2025.

### Quantitative analysis

We used descriptive statistics to characterize survey respondents and disruptions in HIV-related services and clinic operations. Among our full sample of respondents from LMICs, we examined the proportion of clinics that reported providing each service and experiencing disruptions since January 2025 and the extent to which these disruptions were “not fully resolved” by the survey timepoint, stratified by country PEPFAR-support status. We used Chi-squared tests of independence (α=0.05) to examine differences in disruptions between PEPFAR and non-PEPFAR-supported countries. Among the subset of sites reporting disruptions or the introduction of a mitigation measure in response to disruptions, we used Chi-squared tests and Fisher’s exact tests, where indicated by small cell counts (n<5), to examine differences by PEPFAR-support status in the full resolution of disruptions and the continuation of mitigation measures by the survey timepoint. All statistical analyses were performed using SAS 9.4 (SAS Institute, Cary, NC).

### Qualitative analysis

Responses to the survey’s open-ended question were analyzed using a hybrid of deductive and inductive thematic analysis [18–20]. Qualitative data were coded in Dedoose [21] using a primarily deductive approach based on the survey’s domains to elucidate the quantitative results with our qualitative data. Additional inductive codes were created to capture concepts and themes that emerged from participants’ responses but were not represented in the original survey domains.

## III. Results

### Respondent characteristics

Of 174 survey responses received by December 31, 2025, 16 were excluded from the analysis because respondents were in high-income countries and/or left all survey questions unanswered (n=13), or because they appeared to be duplicate responses for the same site/clinic, based on facility name and municipality (n=3). A total of 158 responses from 38 countries were included in our analytic sample, including 123 from 30 countries with PEPFAR support at the beginning of 2025 and 35 from eight non-PEPFAR countries (Table 2). Most respondents in non-PEPFAR countries (31/35, 89%) were in five countries receiving Global Fund support in 2025.

**Table 2.** Characteristics of responding clinics.

|  | **PEPFAR\***<br>**(N=123)** | **Non-PEPFAR‡**<br>**(N=35)** | **Total**<br>**(N=158)** |
| --- | --- | --- | --- |
| **Survey response date**, median (min - max) dates | 11/05/2025<br>(8/26 - 11/07) | 11/04/2025<br>(8/23 - 12/13) | 11/04/2025<br>(8/23 - 12/13) |
| **Facility type** | n (%) | n (%) | n (%) |
| Health center | 27 (22%) | 13 (37%) | 40 (25%) |
| District hospital | 12 (10%) | 1 (3%) | 13 (8%) |
| Regional, provincial or university teaching hospital | 35 (28%) | 2 (6%) | 37 (23%) |
| Dedicated/stand-alone clinic, CBO or drop-in center | 42 (34%) | 19 (54%) | 61 (39%) |
| Multi-site program | 7 (6%) | 0 (0%) | 7 (4%) |
| **Facility administration** |  |  |  |
| Public sector | 58 (47%) | 7 (20%) | 65 (41%) |
| Private/NGO sector | 65 (53%) | 28 (80%) | 93 (59%) |
| **Global Fund support for national HIV response** |  |  |  |
| Global Fund support | 118 (96%) | 31 (89%) | 149 (94%) |
| No Global Fund support | 5 (4%) | 4 (11%) | 9 (6%) |
| **Respondent type** |  |  |  |
| Clinical director/staff | 85 (69%) | 27 (77%) | 112 (71%) |
| Other HIV program staff | 38 (31%) | 8 (23%) | 46 (29%) |
| **Patient volume served** |  |  |  |
| < 50 people | 5 (4%) | 8 (23%) | 13 (8%) |
| 51 - 100 people | 8 (7%) | 8 (23%) | 16 (10%) |
| 101 - 200 people | 10 (8%) | 5 (14%) | 15 (9%) |
| 201 - 500 people | 19 (15%) | 7 (20%) | 26 (16%) |
| 501-1000 people | 14 (11%) | 3 (9%) | 17 (11%) |
| >1000 people | 64 (52%) | 4 (11%) | 68 (43%) |
| Not reported | 3 (2%) | 0 (0%) | 3 (2%) |
| **Patient population served** |  |  |  |
| Predominantly urban | 35 (28%) | 9 (26%) | 44 (28%) |
| Predominantly rural | 16 (13%) | 1 (3%) | 17 (11%) |
| Mixed urban and rural | 72 (59%) | 25 (71%) | 97 (61%) |
| **Patient age-groups served** |  |  |  |
| Children (<10 years) | 75 (61%) | 4 (11%) | 79 (50%) |
| Adolescents (10-19 years) | 94 (76%) | 9 (26%) | 103 (65%) |
| Adults (20+ years) | 123 (100%) | 35 (100%) | 158 (100%) |
| All age groups | 73 (59%) | 3 (9%) | 76 (48%) |

**Abbreviations:** CBO: Community-based organization; NGO: Non-governmental organization; PEPFAR: U.S. President's Emergency Plan for AIDS Relief
**Legend: \*PEPFAR countries:** Burundi, Burkina Faso, Botswana, Cameroon, Democratic Republic of Congo, Colombia, Ghana, India, Jamaica, Kenya, Kyrgyzstan, Cambodia, Laos, Liberia, Lesotho, Myanmar, Mozambique, Malawi, Nigeria, Nepal, Philippines, Rwanda, Senegal, Togo, Thailand, Tanzania, Uganda, South Africa, Zambia, Zimbabwe. **‡non-PEPFAR countries:** Bangladesh, Central African Republic, Fiji, Mexico, Malaysia, Pakistan, Chad, Tunisia.

Most respondents (71%) indicated that they were the clinical director or involved in clinical service provision at their clinic, with the remaining respondents indicating they were involved in programmatic management or program support roles. Overall, 59% of participating clinics were private sector or non-governmental organizations (NGOs), and 41% were public sector clinics, with marked differences between PEPFAR-supported and non-PEPFAR-supported countries; 47% of respondents in PEPFAR-supported countries were based at public sector clinics, compared with 20% of respondents in non-PEFPAR countries.

Respondents represented health centers (25%); district hospitals (8%); referral and/or university teaching hospitals (23%); dedicated HIV clinics, community-based organizations or drop-in centers (39%); and HIV programs comprised of multiple HIV clinics (4%). Respondents in non-PEPFAR-supported countries were predominantly based at health centers and dedicated/stand-alone clinics, with <10% of respondents based at district hospitals or tertiary referral hospitals. Similarly, the majority of respondents in non-PEFPAR countries reported that their clinic served no more than 200 PLHIV, whereas 27% of respondents in PEPFAR-supported countries reported serving a PLHIV population of 201-1000, and 52% reported their clinic or program served more than 1,000 PLHIV.

In both PEPFAR- and non-PEPFAR-supported countries, the majority (61%) of respondents indicated that their clinic or program served a mixed urban and rural population; only 11% reported they served a predominantly rural population. All survey respondents indicated that their clinics served PLHIV aged 20+, and almost half (48%) reported they served PLHIV of all ages (children, adolescents and adults). In PEPFAR-supported countries, a majority of respondents (61%) reported they served pediatric PLHIV (e.g., children 0-9 years), whereas only 11% in non-PEPFAR countries reported serving pediatric PLHIV, with similar differences (76% vs. 26%, respectively) reported for the provision of HIV care to adolescents, aged 10-19.

### Overall disruptions, by setting characteristics

The proportion of clinics reporting any disruptions in the areas of service delivery, medication availability, laboratory services and clinic operations, along with the introduction of selected mitigation measures, are shown in Table 3, stratified by setting characteristics (e.g., PEPFAR vs. non-PEPFAR country support; public vs. private sector administration; facility level; age groups served; and urban vs. rural residence of the population served). Overall, 81% of responding sites reported that at least one HIV-related service had been disrupted or suspended since January 2025 because of changes in U.S. policy or funding. Any disruption of HIV service delivery was more prevalent in PEPFAR-supported countries than non-PEPFAR countries (85% vs. 68%, p=0.03) but did not differ by other setting characteristics.

**Table 3.** Overall status of service delivery disruptions and introduction of mitigation measures, by clinic and setting characteristic.

| Clinic/setting characteristic | Any disruption in... |  |  |  | Introduction of any mitigation measure (n/N) |
| --- | --- | --- | --- | --- | --- |
|  | HIV-related services n/N (%) | Medication shortages or stockouts (n/N) | Laboratory services (n/N) | Clinic operations (n/N) |  |
| All responding clinics | 128/158 (81%) | 112/158 (71%) | 112/158 (71%) | 139/158 (88%) | 123/112 (78%) |
| **Country PEPFAR support** | **p=0.03** | **p<0.01** | **p=0.45** | **p=0.14** | **p=0.05** |
| PEPFAR-supported country | 104/123 (85%) | 94/123 (76%) | 89/123 (72%) | 111/123 (90%) | 100/94 (81%) |
| Non-PEPFAR country | 24/35 (60%) | 18/35 (51%) | 23/35 (66%) | 28/35 (80%) | 23/18 (66%) |
| **Facility administration** | **p=0.49** | **p=0.30** | **p=0.15** | **p=0.93** | **p=0.16** |
| Public sector | 51/65 (79%) | 49/65 (75%) | 42/65 (65%) | 57/65 (88%) | 47/65 (72%) |
| Private/NGO | 77/93 (83%) | 63/93 (68%) | 70/93 (75%) | 82/93 (88%) | 76/93 (82%) |
| **Facility level/type** | **p=0.12** | **p<0.01** | **p=0.14** | **p=0.59** | **p=0.05** |
| Health centers & district hospitals | 47/53 (89%) | 45/53 (85%) | 43/53 (81%) | 46/53 (87%) | 40/53 (76%) |
| Tertiary hospitals | 27/37 (73%) | 24/37 (65%) | 24/37 (65%) | 31/37 (84%) | 24/37 (65%) |
| Dedicated/stand-alone clinics | 47/61 (77%) | 36/61 (59%) | 39/61 (64%) | 55/61 (90%) | 52/61 (85%) |
| Multi-site program | 7/7 (100%) | 7/7 (100%) | 6/7 (86%) | 7/7 (100%) | 7/7 (100%) |
| **Age group(s) served** | **p=0.65** | **p=0.09** | **p=0.37** | **p=0.06** | **p=0.01** |
| Adults only (20+ years) | 44/53 (83%) | 33/53 (62%) | 40/53 (76%) | 43/53 (81%) | 35/53 (66%) |
| All ages (adults, children & adolescents) | 84/105 (80%) | 79/105 (75%) | 72/105 (69%) | 96/105 (91%) | 88/105 (84%) |
| **Population served** | **p=0.49** | **p=0.26** | **p=0.20** | **p=0.64** | **p=0.38** |
| Predominantly urban | 33/44 (75%) | 27/44 (61%) | 27/44 (61%) | 37/44 (84%) | 32/44 (73%) |
| Predominantly rural | 14/17 (82%) | 13/17 (77%) | 14/17 (82%) | 15/17 (88%) | 12/17 (71%) |
| Mixed urban and rural | 81/97 (84%) | 72/97 (74%) | 71/97 (73%) | 87/97 (90%) | 79/97 (81%) |

**Abbreviations:** NGO: Non-governmental organization; PEPFAR: U.S. President's Emergency Plan for AIDS Relief

Similar patterns were observed for reporting of any medication shortage or stockout (71%), which were more prevalent in PEPFAR-supported countries than non-PEPFAR countries (76% vs. 51%, p<0.01). Medication shortages/stockouts were also more prevalent among health centers (85%) and large HIV programs (100%), compared with district and tertiary referral hospitals, with no differences observed across sector or PLHIV populations served. The majority of respondents in both PEPFAR-supported and non-PEPFAR-supported countries reported disruptions in laboratory services (71%) and clinic operations (88%), and differences were not statistically significant for any of the setting characteristics examined.

Reporting of any mitigation measure was high (78%) and more common among clinics in PEPFAR-supported countries (81%) than non-PEPFAR countries (66%). The introduction of any mitigation measure was also more commonly reported by dedicated/stand-alone health centers and large HIV programs (85% and 100%, respectively) compared with health centers (76%) and district or referral hospitals (65%).

### HIV-related services

The largest service disruptions were reported in the areas of pre-exposure prophylaxis (PrEP) (67%), HIV counseling and testing (HCT) (63%) and condom services (64%). Other service delivery disruptions included sexual/gender-based violence (SGBV) services (59%), HIV treatment (56%), contraception/family planning services (47%), PVT services (44%), pediatric HIV services (42%) and tuberculosis or multidrug-resistant tuberculosis (TB/MDR-TB) services (38%).

Disruptions in service delivery and resolution of these disruptions differed by country PEPFAR support status (Figure 1). In PEPFAR-supported countries, 72% of respondents reported disruptions in PrEP consultations, compared with 49% of respondents in non-PEPFAR countries. Additionally, in PEPFAR-supported countries, 65% of respondents reporting disrupted/suspended PrEP services indicated that these disruptions were not fully resolved at the time of the survey, compared with 26% in non-PEPFAR countries. More than half of respondents in PEPFAR-supported countries reported that disruptions in HCT services, SGBV-related services, contraception/family planning services and condoms were not fully resolved at the time of the survey, and more than one-third reported that disruptions in HIV treatment (adult and pediatric) and post-exposure prophylaxis (PEP) were not fully resolved. Additionally, 32% of respondents in PEPFAR-supported countries reported ongoing disruptions in services for PVT and TB/MDR-TB services. In contrast, less than one-third of respondents in non-PEPFAR countries reported ongoing disruptions in HCT, adult HIV treatment, SGBV services, condom services at the time of the survey, and less than one-fifth reported disruptions related to pediatric HIV treatment, PVT, TB/MDR-TB, PEP and contraceptive services that were not fully resolved.

**Figure 1.**
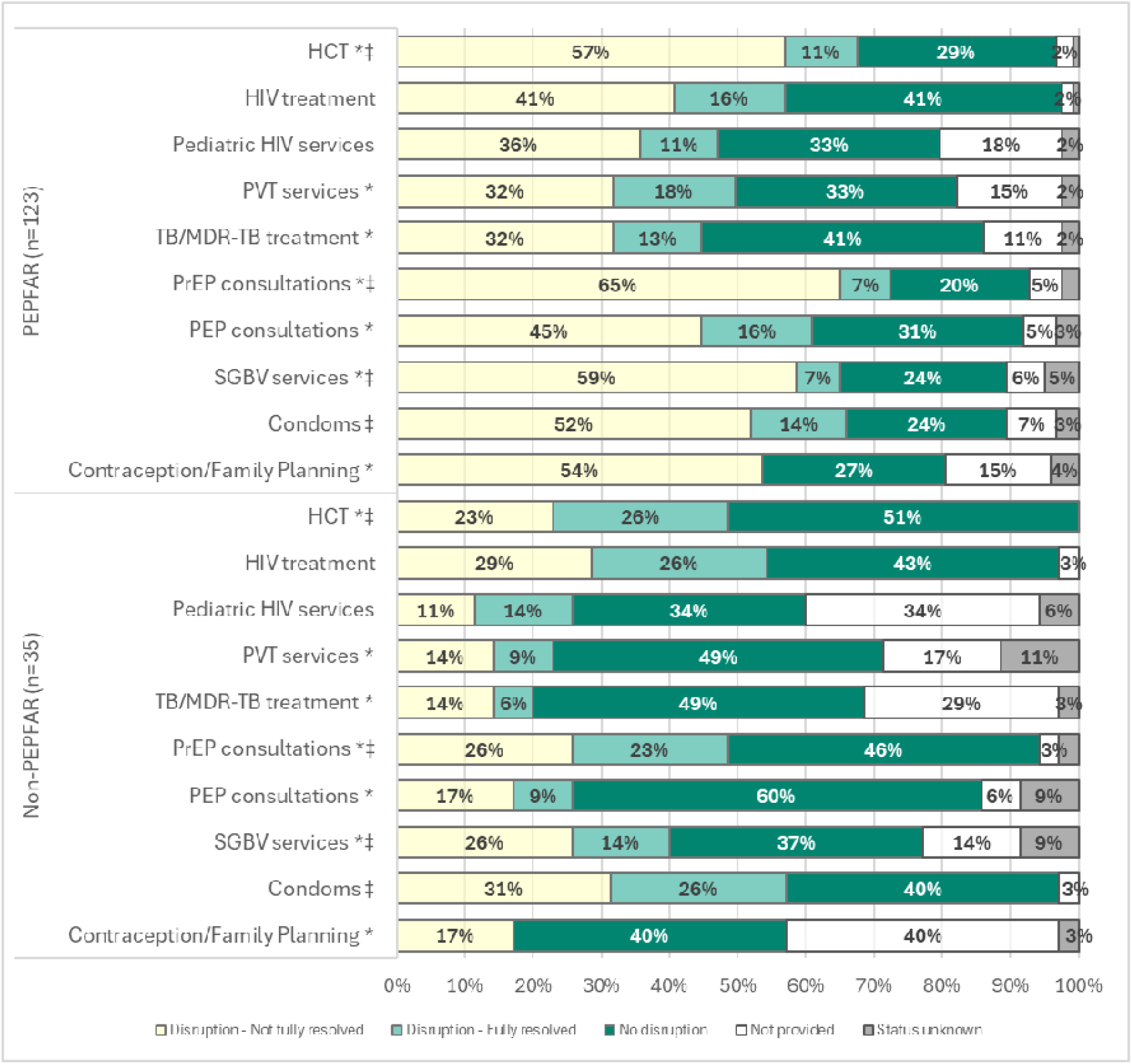
Disruptions in HIV-related services, by PEPFAR country support status. **Abbreviations**: HCT: HIV counseling & testing; PEP: Post-exposure prophylaxis; PEPFAR: U.S. President’s Emergency Plan for AIDS Relief; PrEP: Pre-exposure prophylaxis; PVT: Prevention of vertical transmission; SGBV: Sexual and gender-based violence; TB/MDR-TB: Tuberculosis/Multi-drug resistant tuberculosis. Significant difference between PEPFAR and non-PEPFAR countries (p<.05) in service disruption among clinics that provide the service and reported on disruptions. ‡ Significant difference between PEPFAR and non-PEPFAR countries (p<.05) in resolution of disruptions (fully resolved vs. not fully resolved) among clinics reporting a disruption in services.

### Medication shortages/stockouts

Overall, 51% of respondents reported shortages/stockouts of PrEP/PEP medications, followed by adult antiretroviral (ARV) medications (39%), contraceptives (35%), cotrimoxazole (34%), TB medications (33%), pediatric ARVs (29%) and antimalarials (25%). In PEPFAR-supported countries, 47% and 50% of respondents, respectively, reported that PrEP/PEP and contraceptive shortages were not fully resolved by the survey timepoint, compared with 23% and 11%, respectively, in non-PEPFAR countries (Figure 2). One-quarter to one-third of respondents in PEPFAR-supported countries reported that stockouts or shortages of adult ARVs, anti-TB medications and cotrimoxazole were not fully resolved by the time of the survey, and 20% reported ongoing issues with pediatric ARV availability. In contrast, 11-14% of respondents in non-PEPFAR countries reported ongoing stockouts/shortages for these medications.

**Figure 2.**
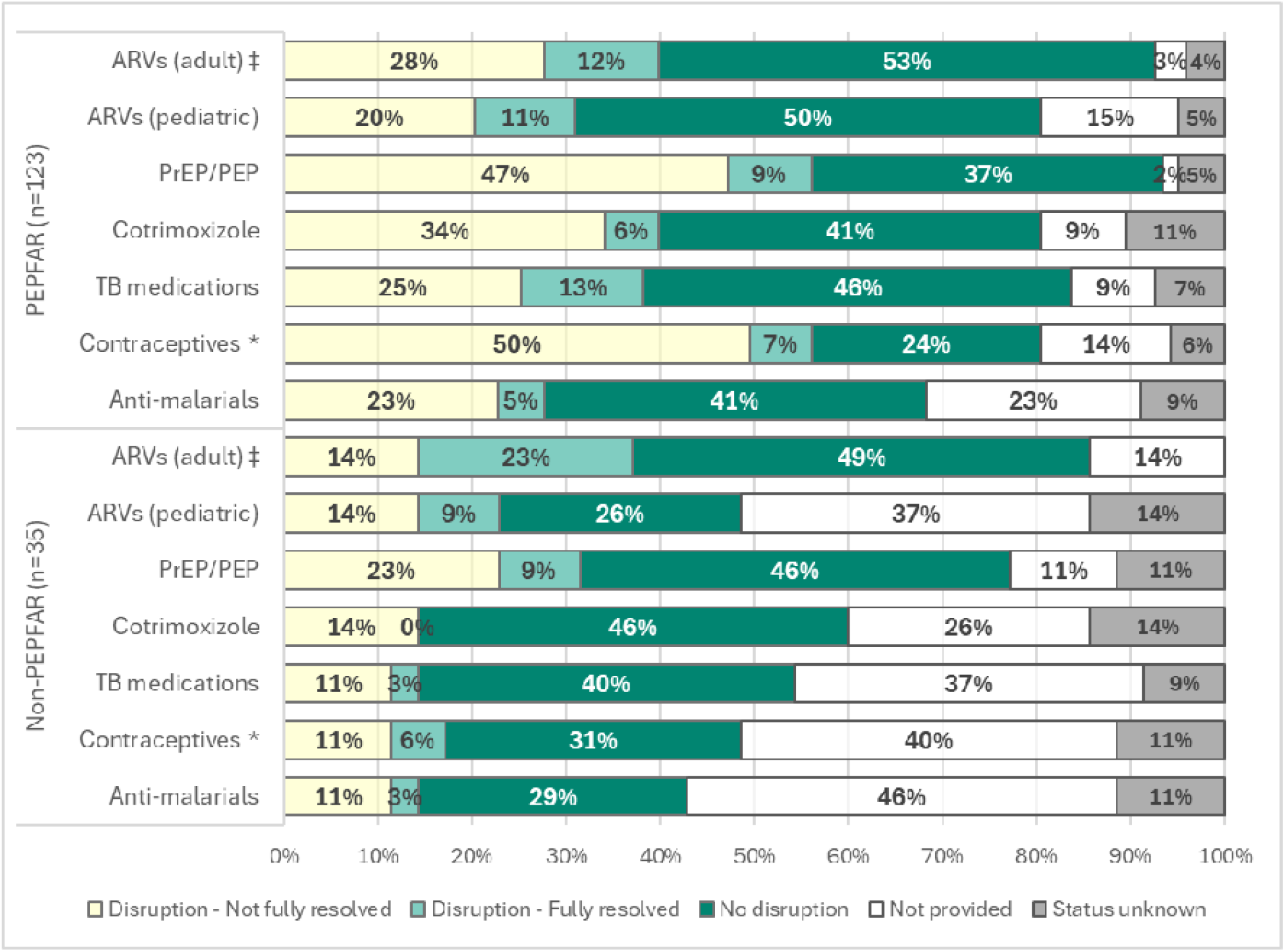
Medication stockouts or shortages, by PEPFAR country support status. **Abbreviations**: ARVs: Antiretrovirals; PEPFAR: U.S. President’s Emergency Plan for AIDS Relief; PrEP/PEP: Pre exposure prophylaxis/post-exposure prophylaxis; TB: Tuberculosis. Significant difference between PEPFAR and non-PEPFAR countries (p<.05) in service disruption among clinics that provide the service and reported on disruptions.

### Laboratory services

Overall, 61% of respondents reported disruptions in HIV testing/diagnosis, followed by disruptions in viral load testing (48%), CD4 testing (41%), early infant diagnosis (EID) (35%), HIV drug resistance testing (35%) and TB diagnosis (34%). Almost half (46%) of respondents in PEPFAR-supported countries reported that disruptions in HIV testing/diagnosis were not fully resolved at the survey timepoint, compared with 29% in non-PEPFAR countries (Figure 3). Additionally, substantial proportions of respondents in PEPFAR-supported countries reported ongoing disruptions in viral load testing (42%), EID (34%), HIV drug-resistance testing (33%), CD4 testing (41%) and TB testing (31%). Fewer respondents in non-PEPFAR countries reported disruptions in laboratory services, and less than one-fifth reported ongoing disruptions in these services at the time of the survey.

**Figure 3.**
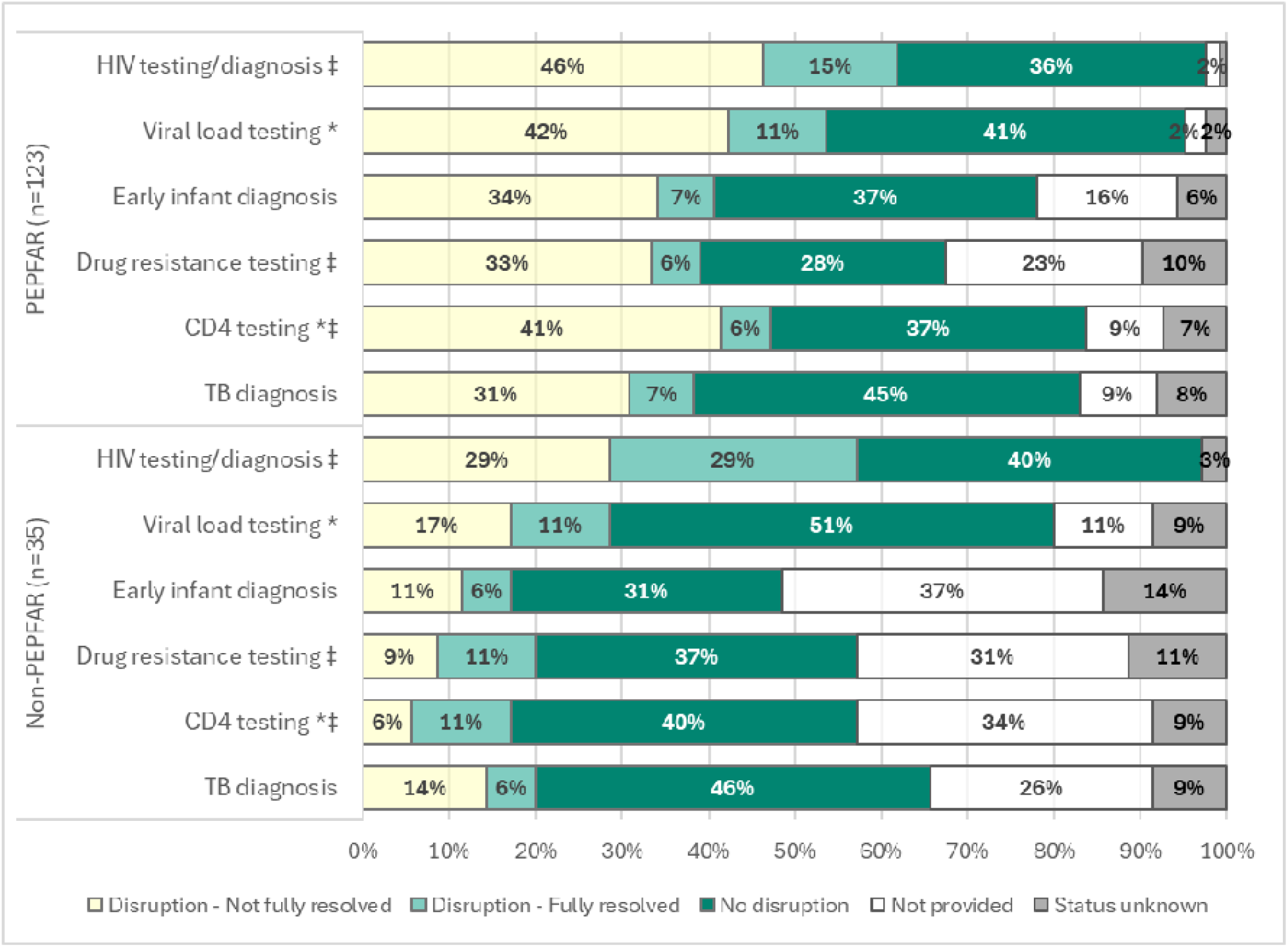
Disruptions in laboratory services, by PEPFAR country support status. **Abbreviations**: ARVs: Antiretrovirals; PEPFAR: U.S. President’s Emergency Plan for AIDS Relief; PrEP/PEP: Pre exposure prophylaxis/post-exposure prophylaxis; TB: Tuberculosis. Significant difference between PEPFAR and non-PEPFAR countries (p<.05) in service disruption among clinics that provide the service and reported on disruptions. ‡ Significant difference between PEPFAR and non-PEPFAR countries (p<.05) in resolution of disruptions (fully resolved vs. not fully resolved) among clinics reporting a disruption in services.

### Clinic staffing and operations

Three-quarters of respondents reported staffing shortages, with 61% reporting salary cuts for HIV providers, 58% reporting interruptions in processing of laboratory specimens, 44% reporting reduced clinic hours or days of operation and 49% reporting interruptions in client record-keeping because of U.S. funding freezes. Other disruptions included community-based activities (68%), tracing clients lost-to-follow-up (67%), adherence support (63%), services for key populations (64%) and suspension of/changes in multi-month dispensing (MMD) of medication (53%). Close to half reported disruptions in other support services, such as TB contact tracing (46%) and services for orphans and vulnerable children (OVCs) (44%).

In PEPFAR-supported countries (Figure 4), the most-affected clinic operations appeared to be PLHIV support services, such as community-based activities (74%), adherence support (72%), client tracing (72%) and services for key populations (68%), along with processing of laboratory specimens (64%). Additionally, more than half of respondents in PEPFAR-supported countries reported other disruptions in clinic operations, including staffing shortages, salary reductions for HIV care providers, reduced hours or days of operation, the suspension of MMD of ARVs because of drug shortages, TB contact tracing, services for OVCs and client record-management. While fewer respondents in non-PEPFAR countries reported disruptions in these areas, close to half reported staff shortages and salary reductions for HIV providers, disruptions in client tracing and adherence support programs and services for key populations. As in PEPFAR-supported countries, relatively few respondents reporting disruptions in clinic operations indicated that these disruptions were fully resolved at the survey timepoint.

**Figure 4.**
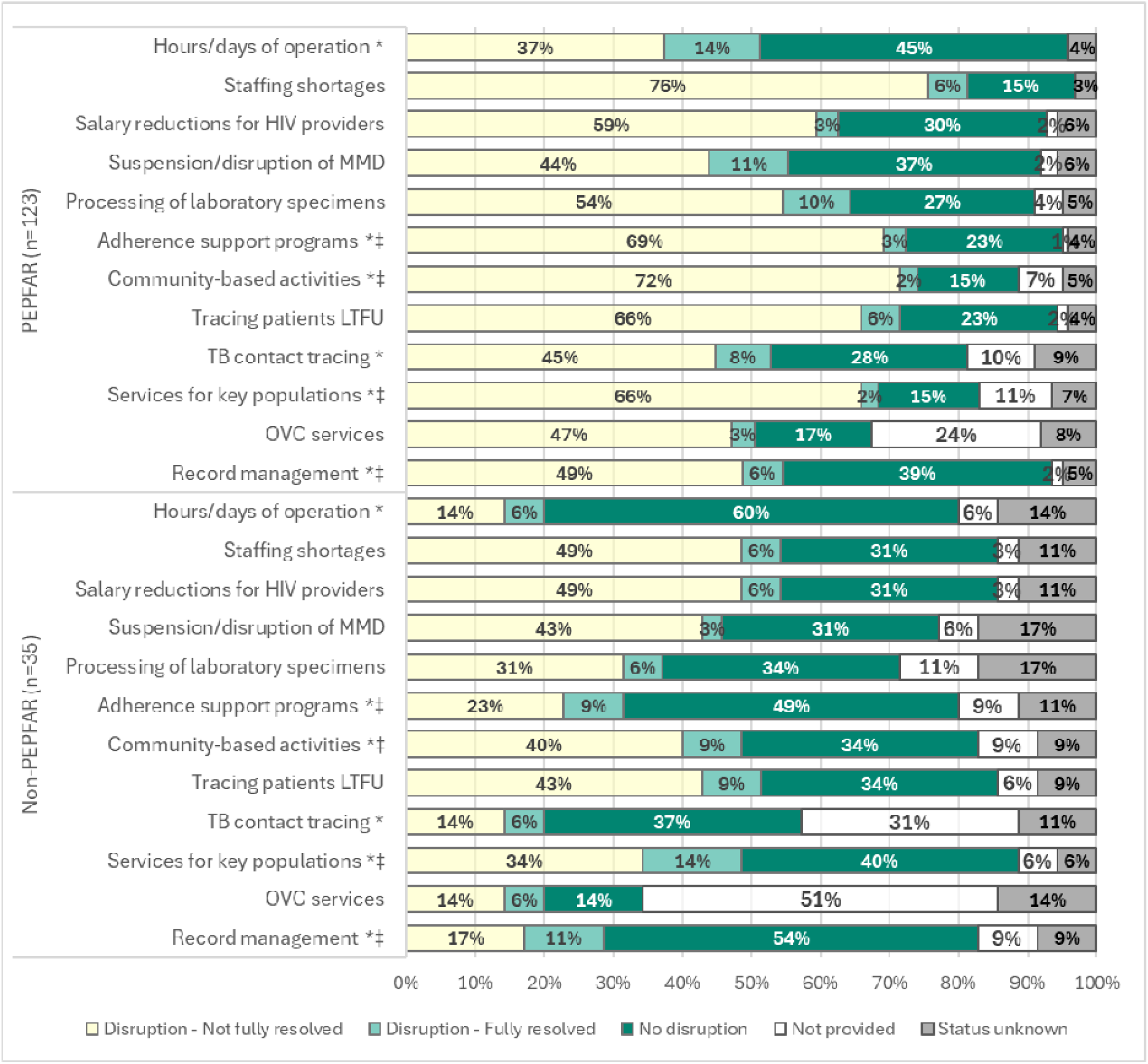
Disruptions in clinic operations and support services, by PEPFAR country support status. **Abbreviations**: LTFU: Lost to follow-up; MMD: Multi-month dispensing of antiretrovirals; OVC: Orphans & vulnerable children; PEPFAR: U.S. President’s Emergency Plan for AIDS Relief; TB: Tuberculosis. Significant difference between PEPFAR and non-PEPFAR countries (p<.05) in service disruption among clinics that provide the service and reported on disruptions. ‡ Significant difference between PEPFAR and non-PEPFAR countries (p<.05) in resolution of disruptions (fully resolved vs. not fully resolved) among clinics reporting a disruption in services.

### Mitigation measures

The most common mitigation strategy reported by respondents was transfer or referral of clients to other clinics, which was reported by 50% of respondents in PEPFAR-supported countries and 43% in non-PEPFAR countries (Figure 5), with 44% and 34%, respectively, indicating that this measure was still in effect at the time of the survey. Other mitigation strategies reported by respondents included advocacy and fundraising efforts (47%), the introduction of MMD of medication (40%), cost-sharing measures (28%) and telemedicine (26%), with these measures being more prevalent in PEPFAR-supported countries than non-PEPFAR countries, and most were still in effect at the time of the survey.

**Figure 5.**
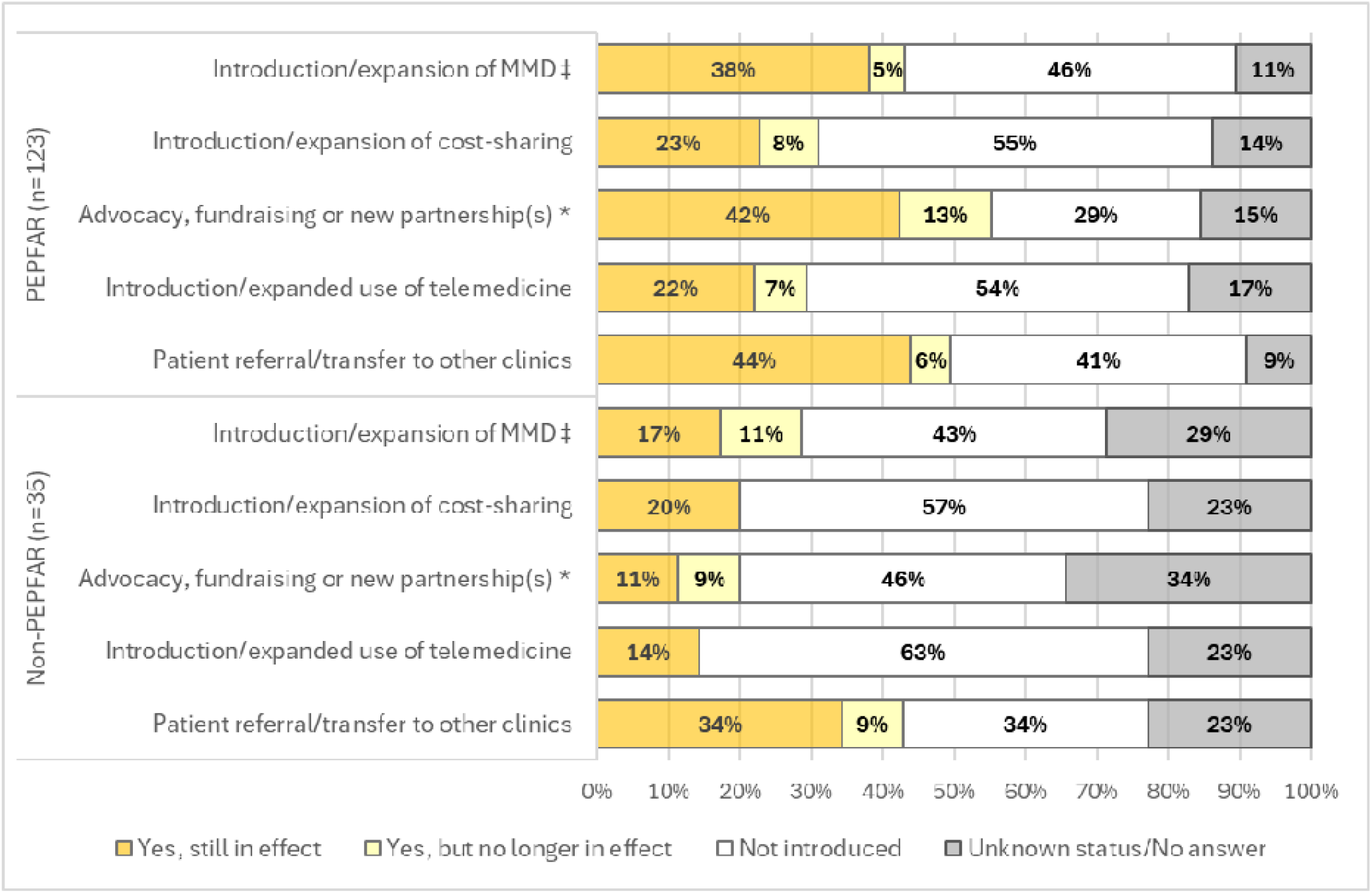
Introduction of selected mitigation measures, by PEPFAR country support status. **Abbreviations**: MMD: Multi-month dispensing of antiretrovirals; PEPFAR: U.S. President’s Emergency Plan for AIDS Relief. Significant difference between PEPFAR and non-PEPFAR countries (p<.05) in the introduction of the mitigation measure (introduced vs. not introduced) among clinics reporting on the mitigation measure. ‡ Significant difference between PEPFAR and non-PEPFAR countries (p<.05) in current status of mitigation measure (in effect vs. no longer in effect) among clinics reporting on current status.

### Qualitative results

Responses to the survey’s open-ended question aligned with our quantitative findings, highlighting disruptions in HIV prevention, HCT, community-based programs and supportive services, such as adherence support and client tracing.

> *“An entire implementing partner programme was shut down that had over 1000 HIV patients being managed, along with HIV Testing, PrEP and contact tracing services being discontinued.”* (Jamaica respondent)
>
> *“Community-based HIV/AIDS prevention and reproductive health campaigns have had to be reorganized or scaled back due to a lack of funding for logistics and community outreach.”* (Chad respondent)
>
> *“Negative impact on patient follow-up. Difficulties in locating patients who have gone missing and delays in antiretroviral treatment and viral load testing.”* (Burundi respondent)
>
> *“There has been drastic reduction in funds for defaulter tracing, psychosocial support and adherence support services.”* (Kenya respondent)

Respondents’ comments also underscored staff layoffs and the resulting consequences for service delivery, along with shortages of commodities and disrupted laboratory services.

> *“A large proportion of staff [were] sent home due to lack of pay.”* (Burundi respondent)
>
> *“The hospital not able to operate the HIV Clinic services at night, during holidays and weekends. The contract staffs reduced by half.”* (Kenya respondent)
>
> *“…several of [our] site offices had to scale down or suspend operations, leading to a reduction in staff and outreach services targeting MSM [men who have sex with men] and MSW [male sex worker] communities…safe community spaces, such as Drop-in Centers that once served as essential hubs for peer support, counselling, and HIV prevention services, have been forced to close in multiple cities due to funding shortages.”* (Pakistan respondent)
>
> *“… as most of the staff have been laid off their duties and working time reduced, work load increased to the remaining staff.”* (Kenya respondent)
>
> *“We have insufficient condoms to distribute and facing a high turnaround time of viral load results.” (*Zimbabwe respondent)
>
> *“Some technical and financial partners linked to U.S. programs (e.g., USAID, PEPFAR, or NGO partners) have reduced their funding or delayed the delivery of medical supplies, HIV tests, and essential medicines.”* (Chad respondent)

Respondents also highlighted the populations most affected by disruptions in HIV programs and services, as well as the population-level impacts they were observing or anticipating.

> *“…We can no longer advertise PrEP nor screen clients for GBV [gender-based violence] even though GBV is rife in Botswana… We are also not allowed to do advocacy work for key populations.”* (Botswana respondent)
>
> *“The impact is particularly felt in already vulnerable communities.”* (Zambia respondent)
>
> *“Global aid cuts are exacerbating already limited access to care, especially for pregnant women, young people, and people living with HIV…”* (Chad respondent)
>
> *“Service deliveries to the rural areas of the country is still not yet back to normal.”* (Kenya respondent)
>
> *“There has been missed opportunities in testing and treatment due to minimal coverage. [We see] increase of new cases of HIV and TB, increase of drug resistance, increase of treatment defaulting, [n]ew cases of vertical transmission in HIV.”* (Kenya respondent)
>
> *“New infections of HIV increasing…. Increased mortality and morbidity rates.”* (Kenya respondent)

In addition to participants’ feedback that was aligned with the survey’s structured questions and deductive codes, inductive coding highlighted the erosion of client trust in the health system and resulting behavioral changes, as well as strains on staff morale and motivation.

> *“Our patients lost all trust in the health system and >70% have dropped out of ART [antiretroviral therapy] and PrEP. When we attempted to refer them to other facilities they indicated that they will take their chances but they don’t trust health systems.”* (South Africa respondent)
>
> “*Poor adherence and so many insecurities cause clients [to] feel we may shut down anytime.”* (Zimbabwe respondent)
>
> *“…In addition to service disruptions, our staff has faced severe challenges due to a 48% salary deduction since January 2025, further impacting morale and sustainability of service delivery. These constraints are not only affecting our ability to provide safe spaces and prevention services but also pose risks to the overall HIV response in Punjab.* (Pakistan respondent)

## IV. Discussion

Aimed at ascertaining the status of HIV care and treatment programs in late 2025, our study documented widespread and ongoing disruptions in various aspects of care that are critical for HIV prevention and management. With responses from clinics and programs across 38 LMICs, our findings highlight the persistence of disruptions identified in stock-taking initiatives and other empirical research conducted in early and mid-2025 [4–11, 22, 23], including life-saving services ostensibly covered under limited waivers granted in February 2025. Our study underscores the breadth of disruptions related to recent changes in U.S. foreign assistance, ranging from routine clinic-based services for adult and pediatric PLHIV to targeted programs, such as community-based services (e.g., ARV distribution) and services for key populations and OVCs, and clinic support functions (e.g., adherence support, client tracing, laboratory processing and record-keeping) that have been critical for reaching HIV treatment goals and sustaining epidemic control in many settings, but are not being prioritized under the new America First Global Health Strategy [24, 25]. Reflecting the far-reaching impacts of changes in U.S. funding, our study also found large disruptions in other essential services, including contraception/family planning, services related to SGBV and malaria treatment—disruptions that modeling studies indicate could result in millions of additional unplanned pregnancies, unsafe abortions and child deaths from causes other than HIV and tuberculosis [26]. Qualitative responses converged with these quantitative findings, underscoring acute on-the-ground consequences of the U.S. foreign assistance freeze, particularly for vulnerable populations, along with concerns not anticipated by our structured survey—concerns, such as the erosion of morale among clinic staff and trust among populations served, that extend beyond immediate resource shortages and which may have long-term effects on the quality, accessibility and utilization of HIV care.

Disruptions in most areas of service delivery, medication availability, laboratory services and other clinic operations and support services were more prevalent among clinics in countries supported by PEPFAR at the start of 2025 than non-PEPFAR countries. Respondents in PEPFAR-supported countries were also more likely to report that disruptions in HIV-related care were not fully resolved at the time of the survey, which was conducted approximately nine months after the U.S. government announced that life-saving services could be resumed under “limited waivers.” Our findings contrast starkly with public statements by U.S. officials in May 2025 that PEPFAR was “85% operative” in terms of delivering HIV care and treatment services [15], and they accord with reports that the Office of Management and Budget (OMB) delayed or avoided disbursing PEPFAR funding approved by Congress in mid-2025 [27–29]. They are also consistent with a recent analysis of PEPFAR monitoring data which showed a 41% decrease in new PrEP initiations for July 1 to September 30, 2025, compared with the same period in 2024, along with decreases in HIV testing, new ART initiations and the numbers served through OVC programs and HIV prevention programs targeting adolescent girls and young women [30].

Our survey also found substantial disruptions in countries not directly supported by PEPFAR, underscoring the reliance on U.S. assistance for HIV and health beyond PEPFAR. Almost half of respondents in non-PEPFAR-supported countries reported disruptions in HCT, HIV treatment, PrEP and condom services, and more than one-third reported stockouts/shortages of ARVs. These findings are consistent with research in South Africa [10] showing that disruptions in HIV-related care in early 2025 extended beyond clinics that were direct recipients of PEPFAR funding. While the extent of disruptions reported in non-PEPFAR countries is surprising, the majority of clinics in these countries (89%) were in countries supported by the Global Fund [17]. With the U.S. being the Global Fund’s largest contributor and providing up to one-third of its funding, U.S. foreign assistance freezes led to substantial cuts to Global Fund country budgets in mid-2025, likely contributing to service delivery disruptions in these settings [29, 31]. Additionally, the five non-PEPFAR countries in our study that received Global Fund support have also historically received U.S. funding to support HIV, family planning and reproductive health, maternal and child health, tuberculosis, nutrition and/or general health services, with reported disbursements for 2025 averaging less than half the disbursements reported for 2024 [15].

While our survey questionnaire was generally aligned with a survey conducted in June-July 2025 among clinics and programs participating in the global IeDEA research collaboration [11], disruptions in HIV care reported via this survey were more extensive than those reported by clinics and programs in IeDEA. For service delivery domains explored in both surveys (e.g., PrEP, HCT, HIV treatment, PVT, pediatric HIV services and TB-MDR TB treatment), disruptions reported in the current survey were more than two times higher than those reported by clinics participating in IeDEA, with larger differences observed in reporting medication shortages/stockouts, disruptions in laboratory services and various clinic operations and support services. The current survey sample reflected a greater proportion of respondents from the private/NGO sector, as well as from stand-alone clinics, community-based organizations and drop-in centers. As such, our survey findings raise concerns that the impacts of U.S. foreign assistance freezes may be more acute among non-governmental partners involved in the HIV response—partners that often serve key populations and vulnerable groups not reached by government health services in many settings because of stigma, lack of privacy and discriminatory treatment of marginalized groups [32–36]. Compared with public sector clinics, these partners may also be more reliant on donor funding to sustain HIV-related services and programs [37, 38].

Most survey respondents reported introducing at least one mitigation measure because of changes in U.S. policy and funding. While more than one-third reported introducing or expanding MMD of ARVs, which could potentially avert treatment interruptions, among those reporting MMD introduction/expansion, more than half reported stockouts or shortages of ARVs because of U.S. funding freezes, with 28% reporting that these stockouts/shortages were not fully resolved by the time of the survey. Almost one-third of respondents reported introducing or expanding cost-sharing measures in response, and these cost-sharing measures largely remained in place among clinics where they had been introduced, reinforcing concerns about treatment access for those who cannot pay for services, medications and essential laboratory testing [39, 40].

Almost half of respondents reported the referral or transfer of clients to other sources of care, with most indicating that this mitigation measure remained in effect at the time of the survey. Past research has shown that program-driven transfers often lead to reduced client satisfaction, interruptions in care and care disengagement [37, 41, 42]. Accordingly, our results raise the specter of treatment interruption and discontinuation for substantial numbers of PLHIV, with qualitative responses indicating that entire programs were shut down and that services for key populations and other vulnerable groups (e.g., young people and pregnant women) were particularly affected. The impacts of drop-in-center closures and service suspensions on key populations may be compounded if government health services deny care to key populations because of discriminatory laws and policies and/or misinterpretation of recent U.S. executive orders and policies related to gender identity and gender-based programing [36]. Marginalized groups may also have more difficulties accessing services where governments are responding to ongoing funding constraints by integrating HIV services into general health services without adequate protections for client privacy and confidentiality [35, 36, 40].

Our findings should be interpreted in the context of several limitations. Designed as an open survey, our study is subject to potential selection and response biases. Few survey responses reflected clinics/programs serving a predominantly rural population, and while one respondent in Kenya noted that service delivery in rural areas had not returned to normal, our survey provides limited insights into the differential impacts of U.S. funding disruptions across rural and urban settings. Additionally, more than half of respondents were affiliated with private/non-governmental clinics—sites that may be better resourced in terms of internet access and linkages to international partners and networks involved in the HIV response. Staff at such clinics may also be more willing to respond to a web-based survey than those working in the public sector, who may not be permitted to report on the situation at their clinic without authorization. It is also possible that staff at clinics experiencing little or no disruptions discounted the relevance of the survey for their setting, or conversely that respondents who were directly affected by salary reductions, increased workload and other negative impacts of the funding freeze illuminated by our survey’s open-ended question were particularly motivated to participate in the survey, In combination, these factors may mean that those most likely to learn about and respond to our survey were those experiencing substantial and sustained disruptions in care because they were more reliant on U.S. funding sources for their services and programs and were not prioritized in governmental mitigation efforts. At the same time, it is also possible that clinics severely impacted by staffing disruptions and other capacity constraints were unable to learn about or complete the survey.

These limitations notwithstanding, our findings align with other empirical research on the impacts of U.S. foreign assistance freezes, while providing an update on the status of HIV-related care in late 2025, and underscoring the extent to which disruptions in service delivery persisted long after the Trump administration’s Executive Orders of January 2025. Additionally, while past research [5, 8, 10] has largely focused on disruptions among PEPFAR-supported clinics and countries, our sample of clinics in 38 countries highlights the broader impacts of recent changes in U.S. policy and funding, which extend beyond PEPFAR-supported countries. A further strength of our study is our integration of quantitative and qualitative findings, which provides additional insights into the impacts of recent funding disruptions on clinics and the populations they serve.

In view of reduced funding commitments from other donor governments [43, 44], and further reductions in and restrictions on U.S. foreign assistance outlined under the “America First Global Health Strategy” [25, 45–47] and the U.S. “Promoting Human Flourishing in Foreign Assistance” Policy [48], the vulnerabilities highlighted by our survey may continue or worsen in 2026 as national responses transition from vertical program models reliant on external funding. Ongoing efforts to monitor the availability and utilization of HIV prevention, care and treatment and mitigate access barriers for hard-to-serve populations will be critical to sustain progress in ending the HIV epidemic.

## Data Availability

All data produced in the present study are available upon reasonable request to the authors

## Authors’ Contributions

EB, FM, and DN conceptualized the study and designed the methodology. EB, DN, FM, and AN contributed to data acquisition. EB performed the statistical analysis, interpreted findings and drafted the manuscript. MK and PK performed the qualitative analysis, and interpreted findings. EB, MK, FM, AN, PK, SND and DN critically reviewed the manuscript for important intellectual content. All authors read and approved the final manuscript.

## Acknowledgements

The authors would like to thank IAS—the International AIDS Society—staff for their help in circulating information about the survey to the IAS membership, as well as colleagues at the World Health Organization, UNAIDS, PEPFAR, amfAR, Medecins San Frontiers, the Georgetown University Center for Innovation in Global Heath and the Elizabeth Glaser Pediatric AIDS Foundation who provided suggestions and help in distributing the survey link to partners.

## References

1. Kates J, Michaud J, Moss K, Dawson L, Rouw A. Overview of President Trump’s Executive Actions on Global Health: KFF; 2026 updated March 31, 2026. Available from: https://www.kff.org/global-health-policy/overview-of-president-trumps-executive-actions-on-global-health/. [Accessed March 31, 2026].

2. KFF. The Status of President Trump’s Pause of Foreign Aid and Implications for PEPFAR and other Global Health Programs 2025 updated February 3, 2025. Available from: https://www.kff.org/policy-watch/the-status-of-president-trumps-pause-of-foreign-aid-and-implications-for-pepfar-and-other-global-health-programs/. [Accessed July 28, 2025].

3. Kates J, Rouw A, Oum S, Wexler A. How Much Global Health Funding Goes Through USAID?: KFF; 2025 updated February 7, 2025. Available from: https://www.kff.org/global-health-policy/how-much-global-health-funding-goes-through-usaid/. [Accessed May 27, 2026].

4. World Health Organization (WHO). The impact of suspensions and reductions in health official development assistance on health systems: Rapid WHO country office stock take 2025. Available from: https://cdn.who.int/media/docs/default-source/integrated-health-services-(ihs)/impact-of-suspensions-and-reductions-in-health-oda-on-health-systems.pdf?sfvrsn=7b1cafcb_7&download=true. [Accessed July 28, 2025].

5. Lankiewicz E, Sharp A, Drake P, Sherwood J, Macharia B, Ighodaro M, et al. Early impacts of the PEPFAR stop-work order: a rapid assessment. J Int AIDS Soc. 2025;28(2):e26423.

6. Global Black Gay Men Connect (GBGMC). Tracking the Freeze: Real-Time Impact on Key Populations 2025. Available from: https://gbgmc.org/impact-us-funding-cuts/. [Accessed July 28, 2025].

7. PrEPWatch. Africa Regional Impacts: Implications of the PEPFAR Stop Work Order 2025. Available from: https://www.prepwatch.org/impact-overview/. [Accessed July 28, 2025].

8. Saito S, Farahani M, Kunda S, Maluantesa L, Guambe A, Worku HA, et al. Effects of Stop-Work orders on HIV testing, treatment and programmes for prevention of vertical transmission in four sub-Saharan African countries. Journal of the International AIDS Society. 2025;28(9):e70034.

9. Mandavilli A. Foreign Aid Freeze Leaves Millions Without H.I.V. Treatment. New York Times. 2025 Feb. 6, 2025;Sect. Section A.

10. Filiatreau LM, Musiello F, Tsagli DD, Chibi B, Mukwekwerere ED, Essack Z, et al. PEPFAR interrupted: real-world consequences of U.S foreign aid instability for HIV service delivery in South Africa. The Lancet Regional Health – Africa. 2026.

11. Brazier E, Duda SN, Ross J, Semeere AS, Tiendrebeogo T, Chimbetete C, et al. Impact of US government funding freezes on the HIV response: findings from a rapid survey in 32 countries. Health Aff Sch. 2026;4(2):qxag020.

12. Younas A, Durante A. Decision tree for identifying pertinent integration procedures and joint displays in mixed methods research. J Adv Nurs. 2023;79(7):2754–69.

13. Harris PA, Taylor R, Minor BL, Elliott V, Fernandez M, O’Neal L, et al. The REDCap consortium: Building an international community of software platform partners. Journal of Biomedical Informatics. 2019;95:103208.

14. Harris PA, Taylor R, Thielke R, Payne J, Gonzalez N, Conde JG. Research electronic data capture (REDCap)—A metadata-driven methodology and workflow process for providing translational research informatics support. Journal of Biomedical Informatics. 2009;42(2):377–81.

15. ForeignAssistance.gov. U.S. Foreign Assistance: Complete dataset: U.S. Department of State on behalf of United States Government agencies reporting foreign assistance.; 2025. Available from: https://foreignassistance.gov. [Accessed March 6, 2026].

16. PEPFAR. PEPFAR Operating Unit (OU) Budgets by Financial Classifications Dataset: PEFPAR; 2025. Available from: https://data.pepfar.gov/datasets#FMD. [Accessed 11/13/2025].

17. The Global Fund. Dataset: Grants disbursements - Implementation Period Currency (2025): The Global Fund; 2025. Available from: https://data-service.theglobalfund.org/downloads. [Accessed 11/13/2025].

18. Proudfoot K. Inductive/Deductive Hybrid Thematic Analysis in Mixed Methods Research. Journal of Mixed Methods Research. 2023;17(3):308–26.

19. Kaur K, Abu-Qamar MZ, Rashidi A, McKay N, Saunders R. Applying Hybrid Coding and Reflexivity in Qualitative Content Analysis: An Exemplar. Glob Qual Nurs Res. 2025;12:23333936251395024.

20. Fereday J, Muir-Cochrane E. Demonstrating Rigor Using Thematic Analysis: A Hybrid Approach of Inductive and Deductive Coding and Theme Development. International Journal of Qualitative Methods. 2006;5(1):80–92.

21. SocioCultural Research Consultants L. Dedoose cloud application for managing, analyzing, and presenting qualitative and mixed method research data. 10.0.59 ed. Los Angeles, CA: SocioCultural Research Consultants, LLC; 2026.

22. Guillén JR, Stevenson M, Talero MÁ B, Torres MA, Wirtz AL. Consequences of United States funding suspensions on community-led HIV services in Latin America and the Caribbean: findings of a rapid service provider survey. J Int AIDS Soc. 2026;29(2):e70081.

23. Zakumumpa H, Adolf A, Kiplagat J, Cherop F, Bosire V, Buregyeya E. Early impacts of PEPFAR funding freeze on HIV service delivery in mid-Western Uganda. BMC Health Serv Res. 2026;26(1).

24. Razavi C, Gaba C, Kates J, Nandakumar A. Which PEPFAR Investments Drive HIV Outcomes? Informing PEPFAR Transition and Scale-Down: KFF; September 2025. Available from: https://www.kff.org/global-health-policy/which-pepfar-investments-drive-hiv-outcomes-informing-pepfar-transition-and-scale-down/. [Accessed January 6, 2026].

25. U.S. Department of State. America First Global Health Strategy 2025. Available from: https://www.state.gov/america-first-global-health-strategy. [Accessed January 6, 2026].

26. Stover J, Sonneveldt E, Tam Y, Horton KC, Phillips AN, Smith J, et al. Effects of reductions in US foreign assistance on HIV, tuberculosis, family planning, and maternal and child health: a modelling study. The Lancet Global health. 2025;13(10):e1669–e80.

27. Gostin LO. The Trump presidency: Cascading global shocks on global health. PLOS Global Public Health. 2025;5(11):e0005385.

28. Kent L. The Trump administration has halted funds for global HIV/AIDS programs. No one knows how big the impact is CNN; 2025. Available from: https://www.cnn.com/2025/09/13/africa/hiv-aids-program-cuts-trump-pepfar-intl. [Accessed April 13, 2026].

29. Partners in Health (PIH). Approved Funding Sits in Banks While Patients Suffer: Partners in Health; 2025 updated September 25, 2025. Available from: https://www.pih.org/article/PEPFAR-Global-Fund-Withheld-Funding-US-Government. [Accessed April 13, 2026].

30. Kates J, Rouw A, Nandakumar A. What We Know from the Latest PEPFAR Data: Analysis of FY 2025 Quarter 4 Results: KFF; 2026 updated Apr 23, 2026. Available from: https://www.kff.org/global-health-policy/what-we-know-from-the-latest-pepfar-data-analysis-of-fy-2025-quarter-4-results/. [Accessed April 27, 2026].

31. Ravelo JL. Devex CheckUp: $1.4B Global Fund cuts hit over 100 countries: devex; 2025 updated July 10, 2025. Available from: https://www.devex.com/news/devex-checkup-1-4b-global-fund-cuts-hit-over-100-countries-110462/amp. [Accessed April 13, 2026].

32. Marano-Lee M, Williams W, Xu S, Andia J, Shapatava E. Contributions of the Community-Based Organization Program Funded by the Centers for Disease Control and Prevention to Linkage to HIV Medical Care. Public Health Rep. 2024;139(6):662–8.

33. Groves AK, Stankard P, Bowler SL, Jamil MS, Gebrekristos LT, Smith PD, et al. A systematic review and meta-analysis of the evidence for community-based HIV testing on men’s engagement in the HIV care cascade. International journal of STD & AIDS. 2022;33(13):1090–105.

34. Joint United Nations Programme on HIV/AIDS (UNAIDS), (PEPFAR) USPsEPfAR. Equity in the HIV response: Assessing progress and charting a way forward Geneva and Washington D.C. 2024. Available from: https://www.unaids.org/sites/default/files/media_asset/equity-in-the-hiv-response_en.pdf#page=44. [Accessed April 14, 2026].

35. Physicians for Human Rights (PHR). “The System is Folding in on Itself”: The Impact of U.S. Global Health Funding Cuts in Kenya 2025. Available from: https://phr.org/wp-content/uploads/2025/07/PHR-Research-Brief-Aid-Cuts-Kenya-2025.pdf. [Accessed April 16, 2026].

36. Physicians for Human Rights (PHR). On the Brink of Catastrophe: U.S. Foreign Aid Disruption to HIV Services in Tanzania and Uganda 2025. Available from: https://phr.org/our-work/resources/on-the-brink-of-catastrophe-u-s-foreign-aid-disruption-to-hiv-services-in-tanzania-and-uganda/. [Accessed April 16, 2026].

37. Wilhelm JA, Paina L, Qiu M, Zakumumpa H, Bennett S. The differential impacts of PEPFAR transition on private for-profit, private not-for-profit and publicly owned health facilities in Uganda. Health Policy Plan. 2020;35(2):133–41.

38. Mandavilli A. New PEPFAR Data Show Worrying Declines in Testing and Treatment for HIV. The New York Times. 2026 April 17, 2026.

39. National Syndemic Diseases Control Council. Kenya Health Service Disruption Assessment: Rapid Results Initiative Report Nairobi, Kenya: National Syndemic Diseases Control Council; 2025. Available from: https://nsdcc.go.ke/wp-content/uploads/2025/09/RRI_FINAL.pdf. [Accessed April 16, 2026].

40. Erica Sedlander, Kyla Lawson, Njeri Wairimu, Kalin Werner, Sharon Mutai, Daniel Mwai, et al. A snapshot of how US funding cuts impacted HIV care in one county of Kenya. Research Square. 2026.

41. Katz IT, Bogart LM, Cloete C, Crankshaw TL, Giddy J, Govender T, et al. Understanding HIV-infected patients’ experiences with PEPFAR-associated transitions at a Centre of Excellence in KwaZulu Natal, South Africa: a qualitative study. AIDS Care. 2015;27(10):1298–303.

42. Cloete C, Regan S, Giddy J, Govender T, Erlwanger A, Gaynes MR, et al. The Linkage Outcomes of a Large-scale, Rapid Transfer of HIV-infected Patients From Hospital-based to Community-based Clinics in South Africa. Open Forum Infectious Diseases. 2014;1(2).

43. Brink Dt, Martin-Hughes R, Bowring AL, Wulan N, Burke K, Tidhar T, et al. Impact of an international HIV funding crisis on HIV infections and mortality in low-income and middle-income countries: a modelling study. The Lancet HIV. 2025;12(5):e346–e54.

44. Global Fund Advocates Network (GFAN). GFAN 2025 Pledge Tracker & Other 8th Replenishment Advocacy Tools 2026. Available from: https://globalfundadvocatesnetwork.org/gfan-2025-pledge-tracker/. [Accessed January 8, 2026].

45. Kates J, Stenoien D, Ruffner M, Nandakumar A. Questions for the America First Global Health Strategy: Transitioning to time-bound, bilateral agreements with partner countries could create gaps in service continuity and health financing: Council on Foreign Relations; November 6, 2025. Available from: https://www.thinkglobalhealth.org/article/questions-for-the-america-first-global-health-strategy. [Accessed January 6, 2026].

46. KFF. KFF Tracker: America First MOU Bilateral Global Health Agreements: KFF; 2026 updated April 8, 2026. Available from: https://www.kff.org/global-health-policy/kff-tracker-america-first-mou-bilateral-global-health-agreements/. [Accessed April 10, 2026].

47. Anya Hirschfeld AK, Thomas J. Bollyky, Stephanie Psaki, Joseph L. Dieleman. Tracking the “America First” Bilateral Health Agreements: Think Global Health; 2026. Available from: https://www.thinkglobalhealth.org/article/tracking-the-america-first-bilateral-health-agreements. [Accessed April 10, 2026].

48. Moss K, Kates J. Understanding the Trump Administration’s “Promoting Human Flourishing in Foreign Assistance” Policy: KFF; 2026. Available from: https://www.kff.org/global-health-policy/the-mexico-city-policy-an-explainer/#cfa66cf9-befc-490d-93fc-b3d26d2cd175. [Accessed March 6, 2026].

